# Vulnerability and informal caregiver: a scoping review

**DOI:** 10.1101/2021.09.02.21263030

**Authors:** Landoni Marta, Petrovic Milica, Ionio Chiara, Gaggioli Andrea

**Affiliations:** CRIdee, Università Cattolica del Sacro Cuore, Milan, Italy; Department of psychology, Università Cattolica del Sacro Cuore, Milan, Italy; ExprienceLab Milan, Italy

**Keywords:** informal caregiver, vulnerability, systematic review, informal care, relationship-based care

## Abstract

**Objective:** This review paper examines the concept of vulnerability in the overall literature and its relation to informal caregivers.

Vulnerability is frequently associated with the sense of being at risk of harm instead of being acknowledged as a human trait to embrace.

Taking a cue from two well-known videos of Brené Brown on how to enact “vulnerability,” we aimed to see if emotional vulnerability - posed/explored as strength or weakness - exists in the informal care context, potentially acting as a powerful resource that teaches individuals to look inward and inspires them to work on themselves.

**Design:** Scoping review

**Data sources:** Following PRISMA-ScR checklist for scoping review, the literature was searched in various databases, including PubMed, Scopus and ScienceDirect.

**Study selection and data extraction:** We systematically searched for: 1) observational studies, experimental studies, and systematic reviews 2) that examined the topic of emotional vulnerability in a caregiving context 3) that were relevant to informal caregivers of older adults 4) that were published from 1976 to 2021 5) in English 6) that included populations ≥18 years old 7) and excluded conceptualization of vulnerability outside of the emotional perspective (i.e., environmental, financial, social, biological, genetic, medical). Two reviewers independently reviewed titles and abstracts, reviewed the full text of relevant articles, and extracted the data

**Results:** From 2502 articles, 21 were determined as eligible.

**Conclusion:** The reviewed articles showed the complexity of the vulnerability construct and several different approaches taken to explore this topic. This research concludes the value of vulnerability for human beings. The paucity of literature on the concept of vulnerability for informal caregivers offers a promising avenue for future research in this field.

**Article summary: Strengths and limitations of this study:**

- This study reviews the conceptualization of vulnerability across literature from 1976 to date, which was never done before
- This study draws a unique parallel between vulnerability in formal care settings and informal care
- This study re-defines the concept of emotional vulnerability in informal care
- The study lacks more concrete first-person perspectives on vulnerability shared by informal caregivers, hence more concrete involvement of informal caregivers would be desirable for representative understanding of the concept of emotional vulnerability in informal care.

## INTRODUCTION

Vulnerability is an essential aspect of human existence that can be addressed through biological definitions, as well as through social and cultural experiences of emotional vulnerability. For instance, the vulnerability to negative emotions has been associated with the individual’s attention bias toward adverse new information [1].On the other hand, several colleagues explore biological vulnerability by defining the trait markers for alcohol and stimulant abuse, addiction relapse, depression, and genetic vulnerability to chronic health issues of mental disorders such as schizophrenia [2, 3].

The vagueness and malleability in the meaning of vulnerability result in a problematic lack of theoretical and analytical clarity of this concept in the mental health context. Adversely, we note an increasingly common use of emotional vulnerability discourses through growing literature across several disciplines and empirical research fields [4-6].

Vulnerability is commonly perceived as an individual weakness, reflecting the openness to both beneficial and harmful influences [7]. Indeed, in the literature, vulnerability has often been defined as a negative state, associated with risk and adversity, harm [8, 9], and inability to protect oneself [10]. A similar approach is presented in the Oxford English Dictionary, where vulnerability is defined as “the quality or state of being exposed to the possibility of being attacked or harmed, either physically or emotionally*”*.

Throughout the literature, the vulnerability concept has been shaped by the factors that potentially stimulate vulnerability in different ways. For instance, environmental, social, psychological and/or physical characteristics. Even though the range of perspectives on vulnerability expanded, healthcare literature still frames this concept by focusing on defining risks and outcomes of vulnerability and maintaining the assumption that vulnerable groups are relatively homogeneous [11].

Following the sparse conceptualization of vulnerability within the health care field, recognising an individual’s interdependence by nature remains generally neglected [12]. In line with that, it can be suggested that each human is receptive to the experience of vulnerability within the context of their lives and open to others’ expression of vulnerability [13]. However, a growing literature poses a more “radical*”* view of vulnerability [14-17], referring to it as the *“*universal vulnerability*”* [18] or the “vulnerability thesis*”* (see Fineman, 2008, 2013). In general, following the universal vulnerability approach, we are all inevitably vulnerable by our human embodiment or “corporality” [14,19].

However, the notion of “corporality” in the health care field shifts towards a “risk-outcome” description of vulnerability by demonstrating how illness could generate vulnerability among patients and caregivers [20]. In essence, it is the awareness of personal physical and emotional vulnerability that helps individuals sympathize with others, imagine the physical pain or understand the emotional responses such as resentment, anger outbursts, or depression to the prolonged course of illness. Studies in the nursing area have demonstrated how nurses strive to provide person-centred care, respect, and support for all patients while creating a safe environment in which vulnerabilities can be revealed, acknowledged and sympathized with [20-22].

The nature of the nurse-patient relationship and the ability to provide person-centred care enables nurses to recognize perceived vulnerabilities and tailor care according to needs, allowing patients to feel less vulnerable. Moreover, a patient’s display of vulnerability is equivalent to a patient who trusts nurses and the broader health care system to heal, restore, and promote their well-being [20]. It is also essential to recognize that, just as patients, nurses can be vulnerable as well. Indeed, nursing work can allow nurses to experience and treasure their sense of vulnerability [21, 23]. Nurses witness profound life and death-related circumstances and are at the forefront of patients lived experiences and stressors. It can be suggested that the very essence of being empathetic can initiate the emergence of vulnerability.

When drawing a parallel between professional nurses and informal caregivers, similar experiences and feelings could be pointed out, arguably differing in intensity. Conversely, only a few studies have explicitly focused on informal caregivers’ vulnerability, with noted studies framing the vulnerability in the negative context (Table 1) rather than posing it as a potentially positive feature that can be used as a tool or path to empowerment.

**Table 1.**
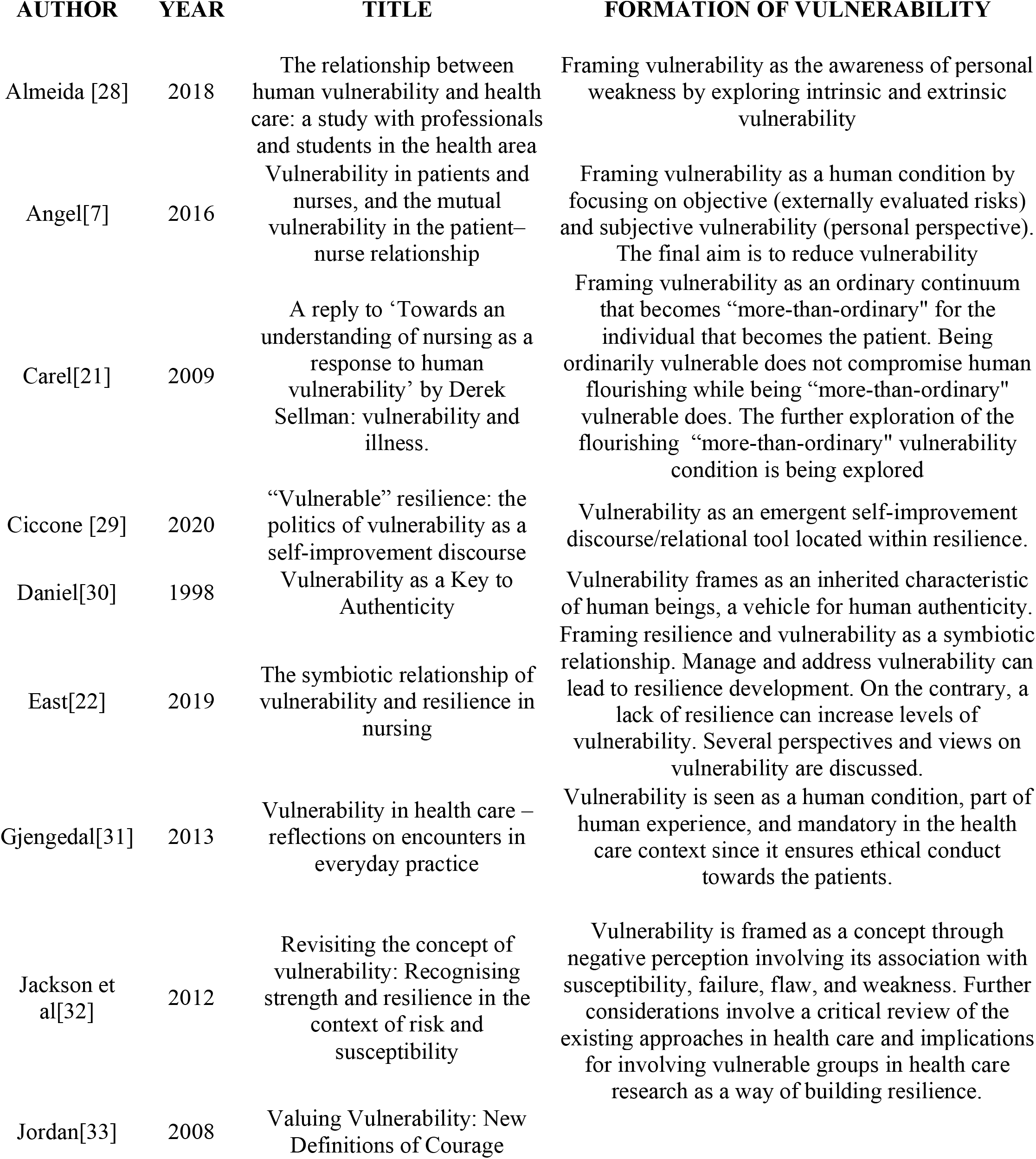
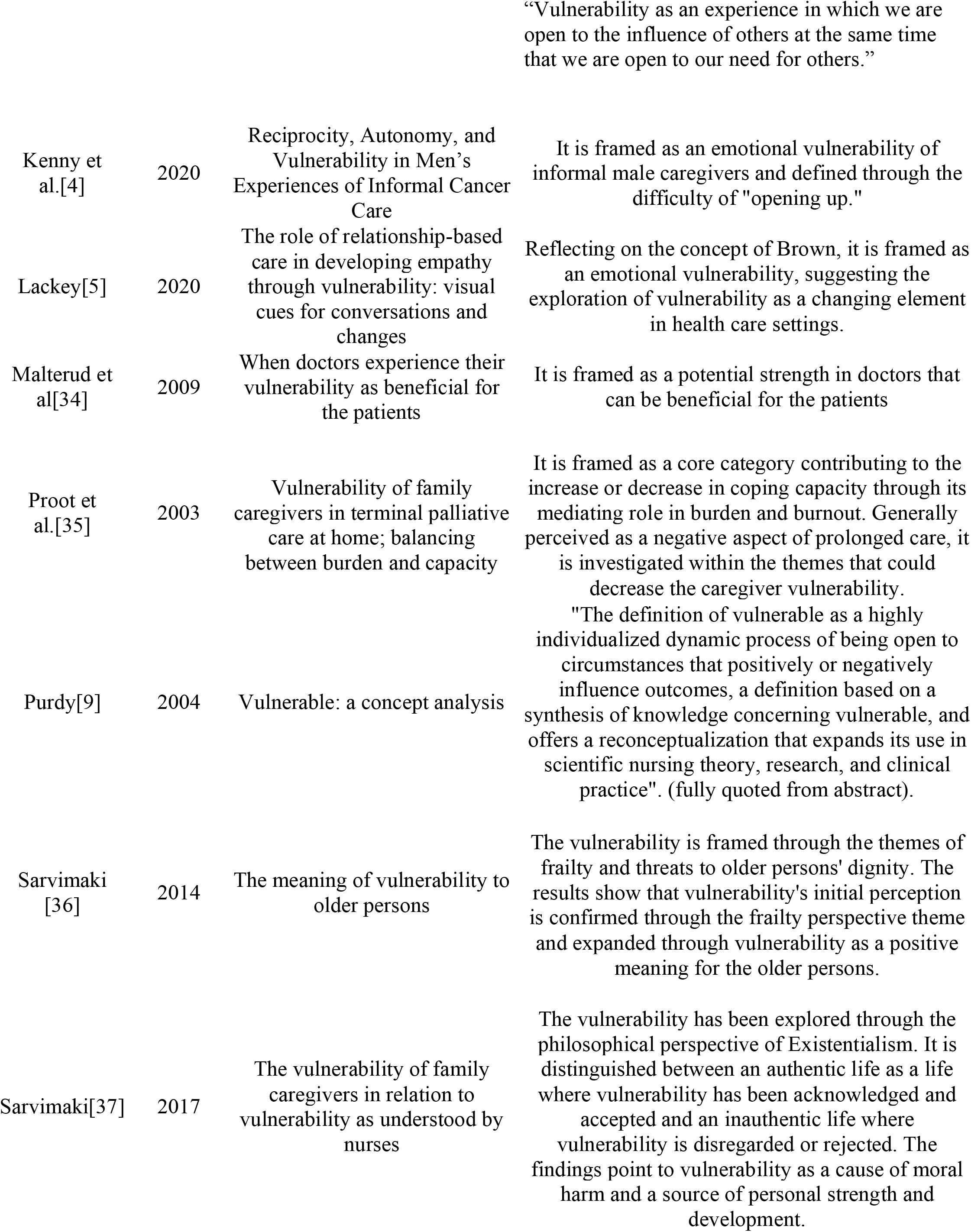
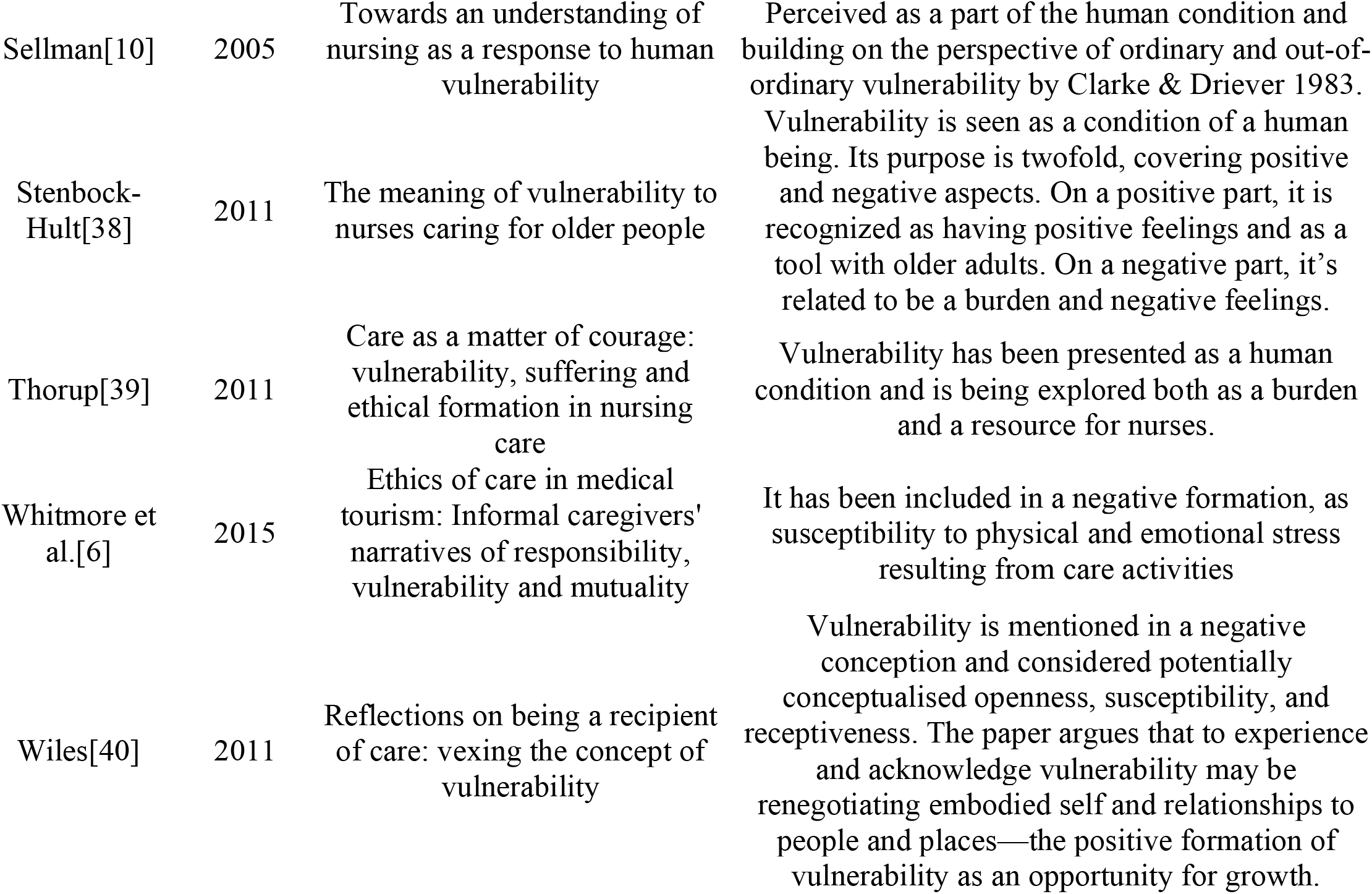
Characteristics of the studies included

In recent years, Brown [24] argued that “Vulnerability is uncertainty, risk, and emotional exposure” in her book “Daring Greatly”. In this context, the vulnerability was defined as the birthplace of courage, creativity and change. Despite widespread interest, there is little empirical evidence to support Brown’s vulnerability theory. Brown [25] further argued that vulnerability is often kept secret, usually due to a feeling of shame. This notion was also expanded, arguing that the extent to which individuals recognize personal vulnerabilities, considering those who are more aware of their vulnerabilities, shows greater resilience to shame. People who do not acknowledge their vulnerability (or perceived invulnerability) as a source of shame, according to Brown, reported more pain and confusion regarding their feelings and the reason behind them. Moreover, Brown [26] noted that strength deriving from vulnerability comes from knowing when and with whom to share the stories of vulnerability to receive empathy in return rather than shame.

Following Brown’s theorization on vulnerability [26] as a source of individual strength, this review investigates vulnerability in the context of informal care, by systematizing the available literature addressing informal caregivers’ vulnerability. Concretely, we aim to explore the existence of emotional vulnerability (i.e., posed/explored as strength or weakness) in the informal care context, to determine the dominant perspective on vulnerability within the field of informal care.

A further aim of this review is to provide directions for potentially harnessing the experience of vulnerability in supporting informal caregivers by identifying current views of informal carer vulnerability, and the potential for shaping the notion of vulnerability in a positive context for informal caregivers.

## METHODS

### Protocol

This review follows the PRISMA-ScR checklist for scoping review, defined by Tricco and colleagues [27]. The checklist contains 27 items that have been completed in this review. The registration of protocol for this scoping review has not been performed.

### Eligibility Criteria

We included 1) observational studies, experimental studies, and systematic reviews 2) that explored the topic of emotional vulnerability in the caregiving context 3) relevant for informal caregivers of older adults 4) published from 1976 until 2021, 5) in English 6) including the population ≥18 years old 7) and excluding conceptualization of vulnerability outside of emotional perspective (i.e., environmental, financial, social, biological, genetic, medical are excluded).

Vulnerability was defined, considering both positive and negative perspectives to ensure the retrieval of all relevant studies inherent to the concept of emotional vulnerability and relevant to informal caregivers. The first definition relied on the dominant perspective of vulnerability as a negative state, associated with risk and adversity, linked with harm [8, 9], and inability to protect oneself [8-10] in the emotional sense. The second definition followed the positive conceptualization of vulnerability by Brown [24] as the birthplace of courage, creativity, and change.

### Information sources

A comprehensive search of electronic databases, including PubMed (1976-2021), Scopus (all years – present), ScienceDirect (all years – present), was conducted. A controlled vocabulary supplemented with keywords was used (Figure 1). Additional search sources included reference lists of the articles, Google Scholar search, and Scimago for verifying the journal ranking of the sources retrieved from Google Scholar. The data search was performed in the period from December 2020 until January 2021. The search strategy included several keywords and three different strings inserted in the various databases such as: “informal care” AND {vulnerability} and “relationship-based care” AND “vulnerability” for Scopus, vulnerability* AND informal care* for Pubmed, informal care AND vulnerability for ScienceDirect (see Figure 1).

**Figure 1.**
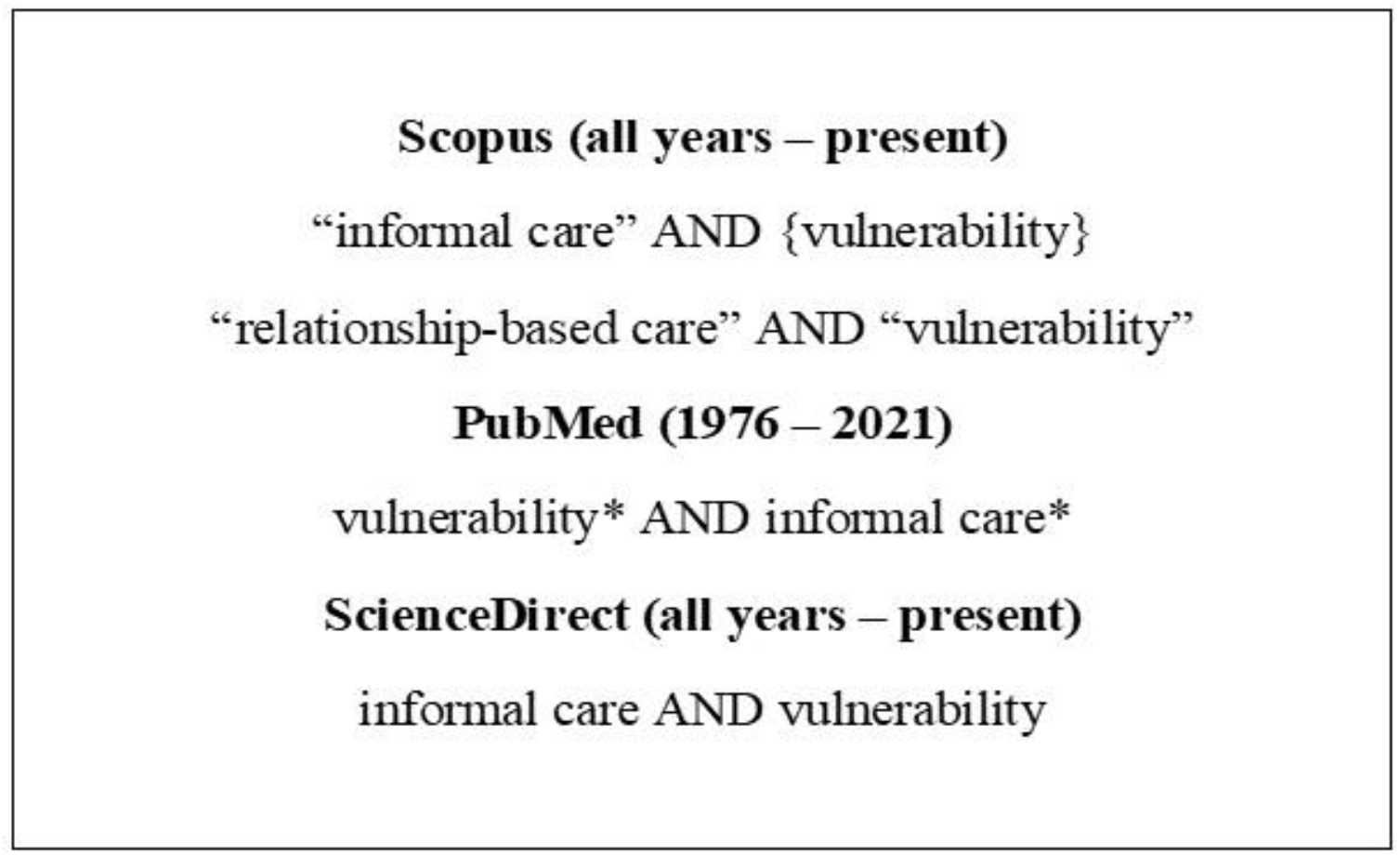
Outline of Keyword Searches Used for This Review

### Selection of sources of evidence

The sources were selected based on the eligibility criteria, with additional assessment against inclusion and exclusion criteria for the abstract/full-text review. The studies that contained the combination of relevant keywords (e.g., including caregiving, care, informal care, and vulnerability) were considered for the abstract/full-text review. Studies selected for abstract/full-text review were screened against the inclusion criteria of (a) studies that were published in academic and peer-reviewed journals, (b) qualitative and quantitative studies (c) that answer “yes” to the following screening questions:

1. Does the study address the concept of emotional vulnerability?
2. Is the study relevant to informal caregivers or conducted in the context of informal care?

The studies that were excluded met the following criteria:

1. Studies addressing vulnerability in the context that is not relevant to informal caregivers or informal care;
2. Studies addressing vulnerability in the context that does not involve emotional vulnerability, but instead economic, medical, social, biological, genetic, cognitive, and environmental definitions of vulnerability;
4. Studies that are not published in English.

### Data charting process

The electronic database search was completed with a total number of 2502 identified articles from Scopus (26), Pubmed (1629) and ScienceDirect (852).

There were two phases of screening: the title screening phase and the abstract/keywords screening phase. Each study was independently checked and double-checked at all stages by the lead author (ML) and the co-author (MP).

Upon completing the screening process, 44 articles were retrieved for the full-text review and sorted with the Mendeley Desktop version 1.19.4 (Figure 2). Eighteen articles were excluded after full text screen, while five retrieved articles were marked as duplicates, leading to the final 21 articles included (Table 1).

**Figure 2.**
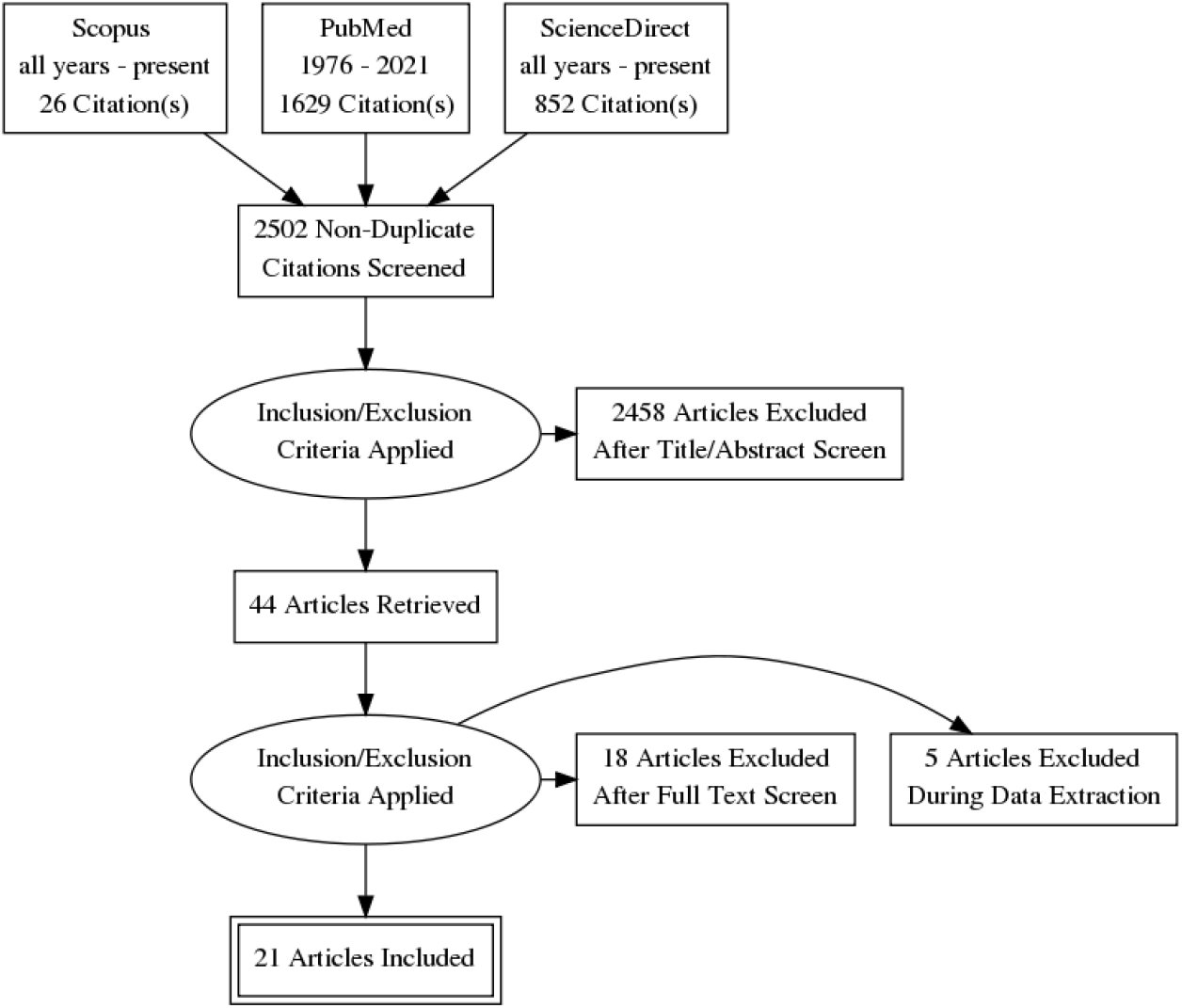
Information flow through different stages of the scoping review process

### Critical appraisal of individual sources of evidence

Using the Critical Appraisal Skills Programme (CASP) Qualitative Research Checklist, the lead author (ML) and co-author (MP) individually and concurrently conducted a quality assessment of each study by running the adapted checklist against the studies included. For instance, this review contains qualitative and quantitative studies as well as critical and perspective papers. Taking into consideration the articles included, the 10-item checklist has been adapted to a 9-item checklist, excluding the seventh item *(“Has the relationship between researcher and participants been adequately addressed”*) and modifying the third item (“*Is the qualitative methodology appropriate*”) since the variety of studies included in this review cannot cover these questions. The critical and perspective articles that cannot be included in any statistical or qualitative analysis but are relevant to the critical discourse have been marked as N/A (Non-Applicable) within the CASP sections that could not be applied to the mentioned studies.

Each question evaluated a specific quality domain such as aim, methodology, recruitment, data, analysis, and findings (Table 2). All points of discord were discussed between the two authors until mutual agreement was achieved.

**Table 2.**
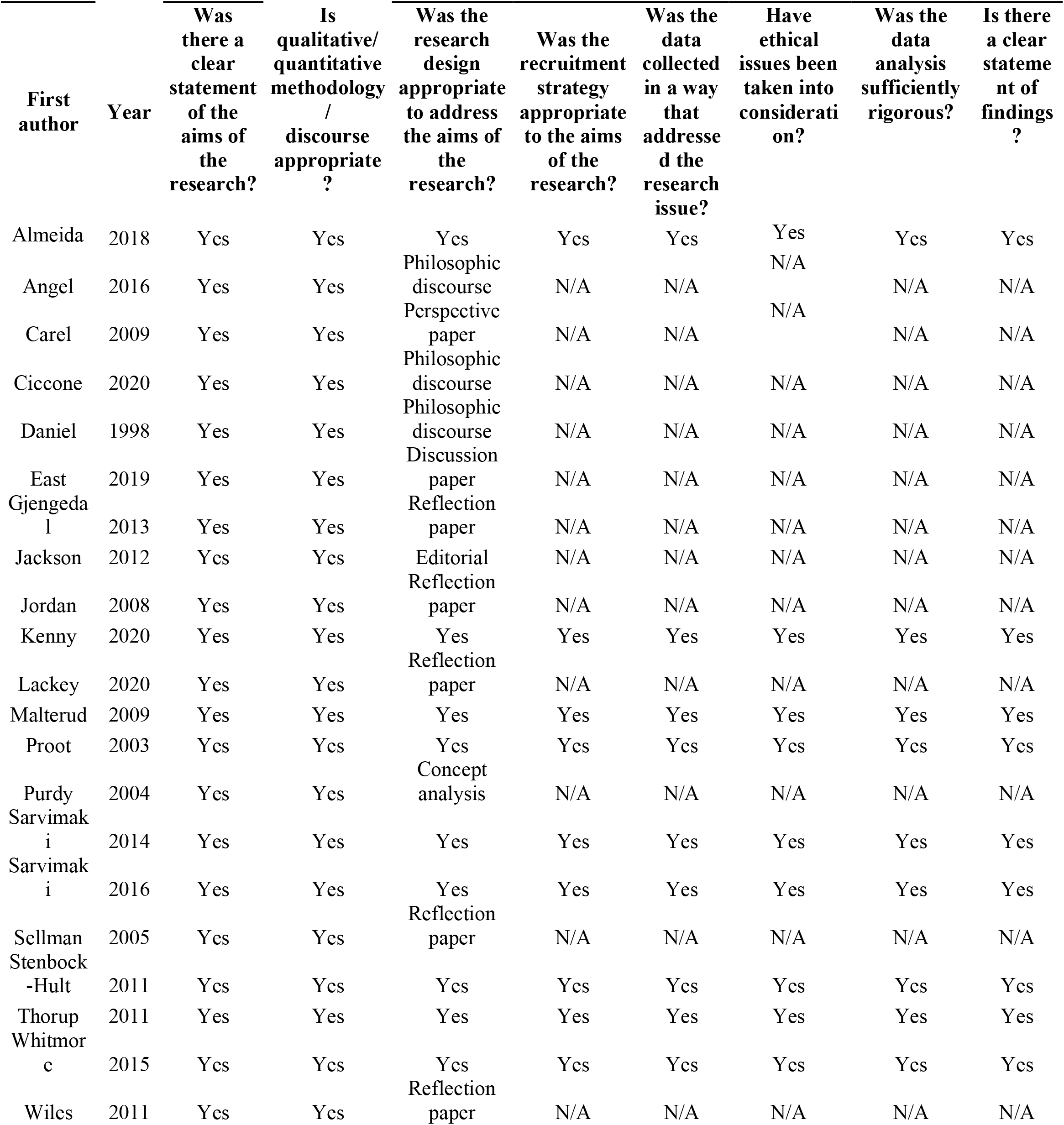
CASP criteria

## RESULTS

We found 2502 manuscripts using our initial search strategy, with 44 manuscripts meeting the inclusion criteria and being selected for the full-text screening after passing the abstract assessment stage. After the full-text screening, we completed the review with 21 studies included with fully quality assessment, data extraction, and coding performed (Figure 2).

Table 1 describes the selected studies and defines the perspective on vulnerability adopted in each study. Table 2 summarizes the included studies’ methodological rigour and quality following the adapted version of CASP the checklist.

Our results reveal the existing complexity of the vulnerability construct and indicate several divergent approaches adopted to investigate this topic (Table 1).

As Figure 3 shows, the core element in most of the studies reviewed is the perception of vulnerability as a human condition. Indeed, as studies indicate, rather than being a trait to avoid, vulnerability was perceived as a trait to embrace, as it allows individuals to celebrate the true essence of what it means to be human. The popular notion of avoiding and protecting oneself from vulnerability is vastly different from perceiving vulnerability as a way of celebrating humanity. Surprisingly, it can be argued that the refusal to experience vulnerability makes us vulnerable [30].

**Figure 3.**
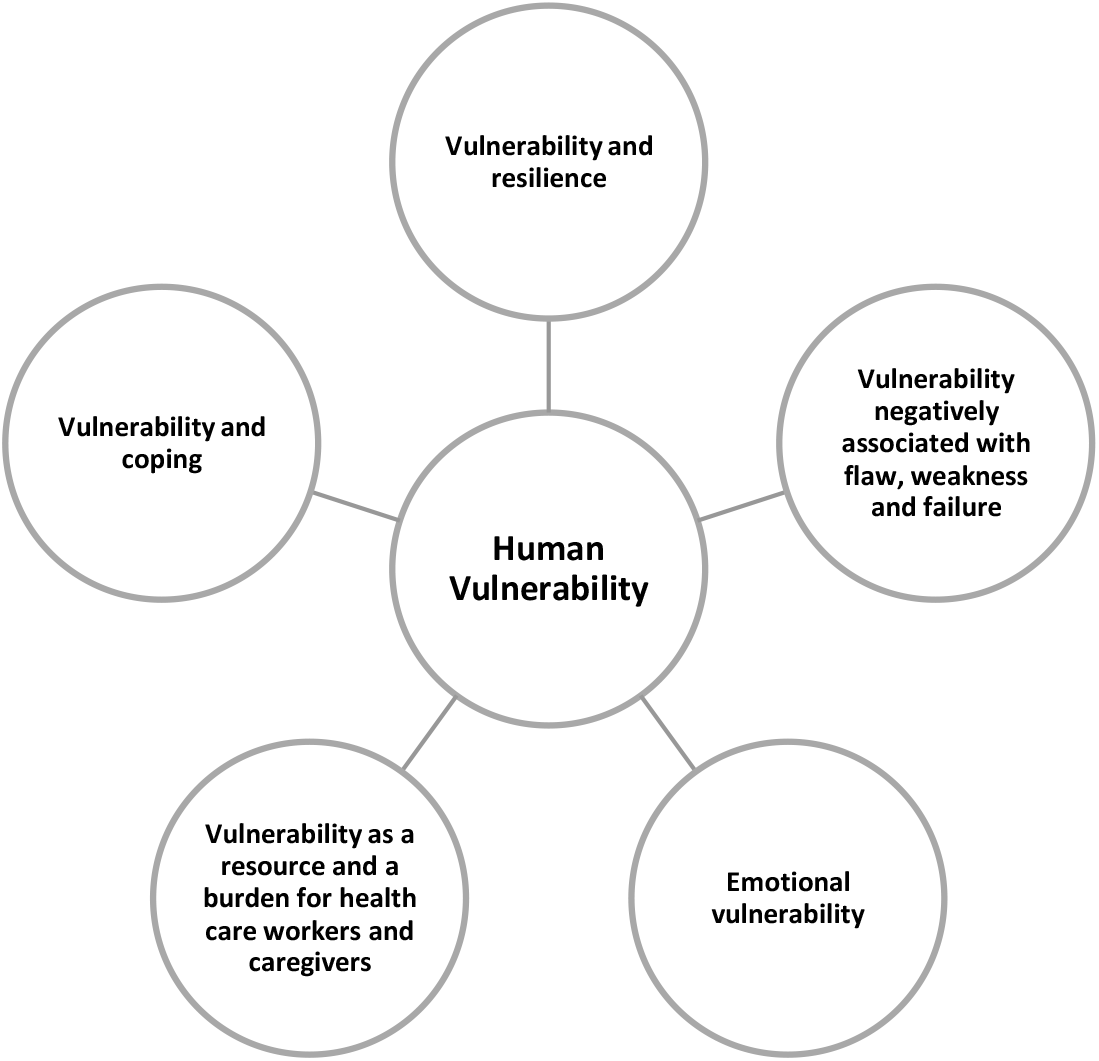
Perceptions of vulnerability

Starting from this core concept, the studies included deepening the exploration by adding several features connected to vulnerability’s central construct, as shown in table 2. However, most of these studies examined the topic in a theoretical way (reflection papers, discussion papers and philosophic discourse papers), while only a few of them in a qualitative way.

Among the qualitative papers, some of them included healthcare workers as a target population [28, 34,38,39], while others included informal caregivers [4,35,37,6 ]. In both groups, vulnerability was perceived as an interchangeable construct of a negative and positive feature. Proot et al. [35], considering vulnerability as a negative construct, identified the influencing aspects increasing (burden, fear, loneliness, lack of support) or decreasing (activities, support, and satisfaction) the experience of vulnerability. Savirmaki et al. [36] identified, through t qualitative analysis, several core themes common for caregivers and nurses, such as: having certain feelings, experiencing moral agony, being emotionally harmed, experiencing courage, protecting oneself, and maturing. These findings are further confirmed by re-emerging in the study of Stenbock-Hult et al. [38]. Therefore, vulnerability as a resource was expressed through the capacity of maturing and developing, having feelings, and courage. On the contrary, vulnerability as a burden was identified in themes such as being harmed and in need of protecting oneself.

However, few studies investigated vulnerability in informal carers, and little knowledge can be found regarding exposure in the informal care context.

## DISCUSSION

This review focused on exploring the concept of vulnerability in the informal care. We scoped the existing literature across three relevant databases that defined emotional vulnerability and its role as either positive or negative experience for informal caregivers. This review aimed to determine how emotional vulnerability is shaped and used in informal care context, potentially leading to a better understanding of the existing perspectives and directing towards positive shift in the definition of emotional vulnerability in the caregiving context. In essence, helping informal caregivers understand, experience, and embrace vulnerability could potentially be the source of empowerment within the caregiving role.

The vulnerability concept stems from the Latin word *vulnerabilis*, signifying wounding or defencelessness against non-physical attack, representing the negative connotation of undesired outcome or an influence. In a widely used definition of vulnerability, the description of the phenomena includes a “dynamic process of openness to circumstances that positively or negatively influence individual outcomes” [7, 9]. Concretely, the openness to any influence is the vulnerability, which is ultimately a path to prosperity and growth but also a potential threat to well-being or infliction of harm by the external environment.

Across the literature, vulnerable populations have always signified specific groups at increased risk of being ill or harmed. Throughout the years, the concept of vulnerability has assumed a negative connotation in Western societies, neglecting the positive aspect of openness to influence that can potentially lead to prosperity and growth [41]. Such representation of vulnerability implies inherent weakness, which is rarely welcomed among individuals since acknowledging vulnerability means accepting to be depicted as dependent, incapable of self-care, or part of a stigmatized group.

However, from a developmental perspective, Daniel [30] and Malone [42] acknowledged how emotional frailty could also be recognized as a positive trait. As an illustration, the quote from the handbook of post-traumatic growth, “I am more vulnerable than I thought but stronger than I ever imagined”, represents the perspective on vulnerability as a source of strength [43]. Similarly, Leffers and colleagues (2004) argued that being threatened can lead to a robust personal response [41], where the perception of being jeopardized or vulnerable leads to the accumulation of power to protect oneself.

Vulnerability could also be defined as “a highly dynamic process of openness to circumstances that positively or negatively influence individual outcomes*”* [9, p. 32]. When one is vulnerable, he/she is susceptible to being “touched*”* by internal and external affective experiences. We honour emotional openness and reward trust in an empathic or compassionate environment. Sarah Lightfoot [44] said, “Making oneself vulnerable is an act of trust and respect, as is receiving and honouring the vulnerability of another”

Few studies focused on the positive aspect of vulnerability aligning with our aim to revise the potential for shaping vulnerability as a positive experience for informal caregivers. The experience of emotional vulnerability within the caregiving role has an early onset. For instance, the vast majority of informal caregivers, providing care for the elderly family member, experience vulnerability at the beginning of the role when the role is either accepted voluntarily or imposed on the caregiver due to the lack of other family member available or willing to provide care.

Furthermore, the onset of the role initiates numerous daily life changes for informal caregiver whose family relationship imminently shifts from the spouse, partner, child, friend into the primary care provider. This role shift implies changes that permanently disrupt the family balance, leaving the caregiver in a state of stress, trauma, and grief. The vulnerability is inevitably experienced but potentially remains hidden since the caregiver becomes the main point of reference for the care recipient and the bridge between the care recipient and the external environment. Hence, requiring a stable mental and emotional appearance since he/she often becomes the spokesperson and the protector of the care recipient.

In literature, vulnerability is also considered an essential aspect of care, considering how being vulnerable could be seen as the fundamental basis for the caring act (e.g., understanding the aggressive phases of some patients with dementia, maintaining the help with hygiene even when the care recipient is no longer aware of his/her needs). In this sense, vulnerability and care are intrinsically linked phenomena.

People rely-on and care for each other because vulnerability is recognized as a universal human condition, also noted as an intrinsic vulnerability in one of the studies included in this review. According to philosophical discourse, the innate nature of being human is related to vulnerability [10]. Anthropological features showed how humans are physically and socially unprepared for adulthood, making them vulnerable [45]. As a result, vulnerability is considered a “condition humana” that concerns everyone [46, p. 461].

Nevertheless, in the care field, the term vulnerable has always been used as a negative concept. Sellman [10] described patients’ vulnerability as “more-than-ordinary” instead of the common vulnerability as a natural human condition that allows flourishing. Physical health problems caused by disease, illness or trauma can lead to increased morbidity or even death, affecting psychological vulnerability and survival [47]. In most of the studies included in this review [6, 7, 28, 32, 35], illness is described through negative aspects (e.g., pain, fear, inability, suffering) and rarely considered as an opportunity to rebuild constructive health habits and use the illness as a tool for self-development to re-establish health, resilience, appreciation for life, and self-respect of personal strength [48, 49, 50].

Within the informal care context, the caregivers often report the sense of loneliness and shame due to sudden or gradual change of the care recipient as an outcome of illness such as stroke or Alzheimer’s disease. Specifically, in the spousal caregiving relations, where the spouse or partner exhibits odd behaviors that he/she cannot control (e.g., uncontrollable shaking, counting repeatedly, word repetition, demonstration of anger by yelling in public places, loss of bowel control), the spouse/caregiver reports feeling ashamed in front of friends and other family members who cannot fully understand the illness and the behavioral outcomes of the illness. Moreover, spousal caregivers often develop a defensive stance towards the external environment, resulting from the need to protect the partner/care recipient who became exposed and vulnerable due to his/her illness. In essence, the spousal caregiver experience vulnerability that they often turn into strength, acting as a protector of the care recipient and their safe connection with the environment.

It is crucial, thus, to recognize the nuanced nature of the relationship between vulnerability and flourishing. According to Sellman [10], the association between the two has a negative correlation: the more vulnerable one is (i.e., “more-than-ordinary”), the less he/she can flourish in life. However, based on recent literature, the relationship appears to be less predictable since everyone is exposed to objective risks and vulnerabilities as human beings in general, and in particular as patients. When illness appears, however, individuals react differently. Since individual reactions to illness differ, there is no necessary link between the disease (or other forms of harm) and personal vulnerability. This is a crucial point for health care providers to consider when interacting with patients: vulnerability is unavoidable and may even be seen as favourable. This aligns with Daniel’s [30]view on vulnerability as a “way of celebrating humanness.*”* Arguably, acts of inhumanity emerge when a person is emotionally invulnerable (e.g., Antisocial Personality Disorder/Psychopathy). Accepting vulnerability is recognizing humanity, which could be considered a “key to authenticity” [30]. It can be suggested that rather than being a trait to avoid, vulnerability is a trait to embrace, as it allows humans to celebrate the true essence of what it means to be human. The popular notion of avoiding and protecting against vulnerability is vastly different from viewing vulnerability as a way of celebrating “humanness”. Surprisingly, it is the abandonment of vulnerability that makes one vulnerable since one becomes vulnerable when they seek to protect from vulnerability by numbing oneself.

However, to perceive vulnerability as a source of strength, one must be aware of his/her vulnerability by recognizing themselves in the other [7]. This is especially important in health care settings, where vulnerability is considered a vehicle for authentic nursing practice. Vulnerable people require care, and caregivers reach out to provide such care.

Vulnerability is the first level of caring assessment, according to Gadow [51], since it is the most urgent one. “For the sick person, illness becomes a situation of limitless vulnerability” [51, p.4] that is often well understood by healthcare providers. Concretely, the exposure to others’ vulnerability rouses personal feelings of vulnerability [28], while the healthcare providers, same as informal caregivers, confront others’ vulnerability daily. In line with that, it can be suggested that the presence of a care recipient’s vulnerability, in turn, provokes the vulnerability of an informal caregiver.

It could be suggested that a person who negatively copes with his/her vulnerability or of the others could experience feelings of maladjustments such as social isolation, burden, depression, and stress. Otherwise, a person who is willing to accept, embrace, and be compassionated with his/her vulnerabilities or that of others, would cope better alone and with people. Indeed, as argued by Brown [24], self-compassion has three parts: recognizing and opening oneself with awareness to the emotional pain; reminding oneself that suffering is an ordinary reality for everyone and that one does not need to feel ashamed or isolated; responding with loving kindness and not self-criticism. Following this argument, nurses and caregivers can choose to form relations where they have *“*power with” or “power over” the care recipient.

The integrity of power, the ability to accomplish something, is preserved through “power with” relationships, maintaining equity of authority. “Power with” relations, according to Watson [52], are those in which “we learn how to be human from one another by identifying ourselves with others or finding their dilemmas in ourselves” (p.59). Vulnerability facilitates individual authenticity. Nurses or caregivers become authentic when they engage in a mutual position of power - when they use “power with” rather than “power over” for forming relations.

As some studies suggested, vulnerability occurs in relationships where people are given the care to explore their full range of emotions in a safe and mutual environment. As Jordan [33] argued, “It is about cultivating connection and to go a step further, I believe it is also about cultivating courage in connection. This helps us cope, stay in our vulnerability, and feel like we are a part of something bigger than our own fears.”

Considering Brene’ Brown’s work on vulnerability [24,53,54] as a core of complex emotions—it’s recognizable how there is vulnerability without empathy [24].

Malterud and colleagues (2009), for example, found empathy as a fundamental component of a physician’s vulnerability. By investigating instances in which physicians’ sense of vulnerability was beneficial for the patient, they discovered how physicians would identify more closely with the patient’s perception of the problem after recognizing similarities between their own lives and the patient’s story [34].

Similarly, in Stenbock-Hult’s study [38], the vulnerability was conceived as a positive development. Being human was identified as the core meaning of vulnerability, encompassing all other connotations: openness, participation, awareness, and sensitivity [55]. It also entailed taking responsibility for oneself and others and having the courage to do so [55]. When vulnerability was combined with courage, it was seen as a strength and a resource. In contrast, when vulnerability was not connected with courage, it was seen as a liability and a weakness. Daring to confront and express one’s feelings and encountering vulnerable older people required courage. It was thought that having the courage to face their vulnerability was a requirement for providing good care. Courage meant admitting their limitations as well as confronting their vulnerability [38].

Furthermore, in a study by Thorup and colleagues [39], it has been demonstrated that assisting patients in confronting their vulnerability and suffering required nurses’ courage that was present in those moments. The nurses reported that they would be unable to predict and mitigate the patient’s reaction in challenging situations if they lacked courage. Consequentially the nurse must accept uncertainty about the patient’s response or have the courage to venture into unknown territory.

In line with that, Arman and colleagues note that courage in health care is defined as “a nurse’s willingness to accompany patients on their journey to overcome their suffering, no matter where the road leads”. This journey is unpredictable, and it appears to require the willingness and ability to witness patients’ vulnerability and suffering. Similarly, Almeida and colleagues [28] suggested that accepting vulnerability as an inseparable part of human reality, in turn, increased the value of caring among caregivers.

Finally, also in Sarvimaki [36], the common core theme was being human. Even though nurses could be considered professional caregivers, on the contrary to family caregivers, both types of care are based on a human-to-human relationship. The family caregivers, as nurses, demonstrated authenticity by expressing how this relationship evoked feelings and caused moral agony and harm. They were conscious of their responsibilities as human beings and dared to face the pain associated with living authentically. They also demonstrated the courage and commitment to living an authentic life by admitting their flaws and speaking out against wrongdoing. The family caregivers gained confidence and strength as they became aware of their vulnerability, realized they had a meaningful task and realized they could manage things despite the situation’s vulnerability. Strength and satisfaction came from a close relationship with the person being cared for [36].

The importance of relationship-based care (RBC) is well described in the research of Lackey [5]. The author strengthened the importance of 3 main areas for care: self-care, care for colleagues and care for patients and families.

Focusing on the first RBC relationship, self-care, can alleviate the fear of vulnerability. By bringing awareness into the equation and gently examining the behaviours we see in ourselves, self-reflection breaks the cycle of internal drama that can occur in the busy healthcare workplace [56]. We can start by asking ourselves, “Who am I?” “allowing us to check in with our current state, give space to what we are going through, and lead us to a sense of strength and courage in our self-care. As a result, the anxiety associated with vulnerability may be reduced. We intentionally work on ways to foster healthy connection and communication as we face the ever-changing challenges of our work; we engage in shared decision-making as we face the ever-changing challenges of our work, and we celebrate and openly thank each other in caring for colleagues in the second RBC relationship.

The third caring relationship, care for patients and families, requires intentionality to shape a caring culture, just like the other RBC relationships. We must learn to be present in every encounter and resolve to establish the connection to function within the openhearted uncertainty of vulnerability. Maintaining belief, knowing, being with, enabling/informing, and doing for [57] the five caring processes described by Kristen Swanson—ensure that our behaviours communicate caring while providing us with evidence-based tools guide our interactions with patients [58].

“In a culture where people are afraid to be vulnerable, there can be no empathy” [26].

We must look beyond traditional approaches to embrace uncertainty. The three RBC relationships provide a framework for exploring vulnerability, allowing the principles and practices to open our hearts and lead us to the genuine empathy we seek when serving our patients, ourselves, and each other.

### Limitations

The evident literature gap in the concept of vulnerability in the informal care context limited the range of studies available for this scoping review. Further quantitative data is required for more concrete findings and the development of the measures focused on assessing emotional vulnerability. Current literature, including the perspective papers and the philosophical discourse, approach the concept of vulnerability with no concrete measures or guidelines of quantifying the emotional vulnerability. Therefore, the scientific rigour of the included studies could not be thoroughly assessed with the existing checklists since the included studies did not fit the clear criteria for qualitative or quantitative research.

## CONCLUSIONS

The sparse conceptualization of vulnerability is also reflected in the informal care field. The lack of literature addressing the concept of vulnerability for informal caregivers provides a valuable path for future research directions within this field. Informal caregivers are confronted with vulnerability early on in the role through emotional and actual life changes in a personal relationship with the care recipient and other family members involved in care. It can be suggested that the concept of vulnerability is not well understood nor promoted enough to be acknowledged but is felt in a reported sense of isolation, helplessness, feeling exposed to the social environment, to the medical setting, feeling misunderstood, and burdened. The evident experience of vulnerability in informal care can be utilized as a potential tool for facilitating informal caregivers’ adaptation to the assumption of the role and the life changes that occur upon assuming the position. Enabling and empowering informal caregivers to recognize, acknowledge, and accept the experience of vulnerability towards the care recipient, the care recipient, and the caregiving circumstances, allows the utilization of the vulnerability as a tool. Finally, acknowledging the experience of vulnerability creates a space for flourishing, potentially leading to the improved health and wellbeing of informal caregivers within the informal care context.

## Data Availability

All data relevant to the study are included in the article

## Author Contributions

LM and MP conceptualised and designed the study, designed the data collection instruments, collected data, carried out the initial analyses and interpreted data, drafted the initial manuscript, and reviewed and revised the manuscript. IC and GA supervised data collection and critically reviewed the manuscript for important intellectual content. All authors approved the final manuscript as submitted.

## Patient consent for publication

Not required.

## Data availability statement

All data relevant to the study are included in the article

